# A Longitudinal Study of Executive Function in Daily Life in Male Fragile X Premutation Carriers and Association with FXTAS Conversion

**DOI:** 10.1101/2023.08.31.23294855

**Authors:** David Hessl, Karina Mandujano Rojas, Emilio Ferrer, Glenda Espinal, Jessica Famula, Andrea Schneider, Randi Hagerman, Flora Tassone, Susan M. Rivera

**Affiliations:** MIND Institute, University of California Davis Health, Sacramento, CA, USA; Department of Psychiatry and Behavioral Sciences, University of California Davis School of Medicine, Sacramento, CA, USA; Department of Psychology, University of California Davis, Davis, CA, USA; Department of Pediatrics, University of California Davis School of Medicine, Sacramento, CA, USA; Department of Biochemistry and Molecular Medicine, University of California Davis School of Medicine, Davis, California, USA; Center for Mind and Brain, University of California Davis, Davis, California, USA; Family Caregiving Institute, Betty Irene Moore School of Nursing, University of California Davis, Sacramento, California, USA; Department of Psychology, University of Maryland, College Park, Maryland, USA

**Keywords:** *FMR1* gene, fragile x-associated tremor/ataxia syndrome, neurodegeneration, fragile X, CGG-repeat, trinucleotide repeat, executive dysfunction, working memory, attention, inhibitory control

## Abstract

**Background:** Men with fragile X-associated tremor/ataxia syndrome (FXTAS) often develop executive dysfunction, characterized by disinhibition, frontal dyscontrol of movement, and working memory and attention changes. Although cross-sectional studies have suggested that earlier executive function changes may precede FXTAS, the lack of longitudinal studies have made it difficult to address this hypothesis.

**Methods:** This study included 66 FMR1 premutation carriers (PC) ranging from 40-78 years (Mean=59.5) and 31 well-matched healthy controls (HC) ages 40-75 (Mean 57.7) at baseline. Eighty-four participants returned for 2-5 follow up visits over a duration of 1 to 9 years (Mean=4.6); 28 of the PC developed FXTAS. The Behavior Rating Inventory of Executive Function-Adult Version (BRIEF-A) was completed by participants and their spouses/partners at each visit.

**Results:** Longitudinal mixed model regression analyses showed a greater decline with age in PC compared to HC on the Metacognition Index (MI; self-initiation, working memory, organization, task monitoring). Conversion to FXTAS was associated with worsening MI and Behavioral Regulation Index (BRI; inhibition, flexibility, emotion modulation). For spouse/partner report, FXTAS conversion was associated with worsening MI. Finally, BRIEF-A executive function problems at baseline significantly predicted later development of FXTAS.

**Conclusions:** These findings suggest that executive function changes represent a prodrome of the later movement disorder.

## 1. Introduction

The Fragile X Messenger Ribonucleoprotein 1 (FMR1) premutation is a trinucleotide (CGG) expansion that occurs in approximately 1 in 110-250 females and 1 in 400-850 males worldwide **[1,2]**. Premutation carriers (PC) are at increased risk of developing a neurodegenerative disease, fragile X-associated tremor ataxia syndrome (FXTAS), characterized primarily by tremor and/or cerebellar gait ataxia, and often with co-occurring conditions such as peripheral neuropathy, parkinsonism, executive dysfunction, and dementia. FXTAS is accompanied by brain white matter changes (T2 hyperintensities in the cerebellum and cerebrum), general brain atrophy, and ubiquitin-positive intranuclear inclusions seen in post-mortem neuropathology studies.

Men with FXTAS often experience executive dysfunction (ED) during the progression of the disease, which is a minor criterion for FXTAS diagnosis. ED in FXTAS often includes disinhibition, frontal dyscontrol of movement, working memory loss and/or deterioration of attention. ED in FXTAS was initially recognized through examination of a series of cases utilizing psychiatric assessment **[3]**, but it was not until the work of Jim Grigsby and colleagues that a clearer picture of the cognitive phenotype was appreciated through standardized neuropsychological assessments and specialized tests that measure the frontal/executive control of movement **[4–6]**. These studies revealed that while verbal intelligence and domains of perceptual reasoning not involving motor coordination are relatively spared, measures tapping regulation of manual motor movements, verbal fluency, general mental status, information processing speed, temporal sequencing, working memory, inhibition, short-term memory, and cognitive flexibility tended to show significant deficits. This study’s early focus on male PC who were asymptomatic for FXTAS also introduced an important question for the field: Do carriers without FXTAS demonstrate executive dysfunction and if so, are they stable traits associated with the premutation, or do they represent a prodrome of the movement disorder?

Initial clues to begin to answer this question came soon in the form of a cross-sectional study of PC males ranging in age from 18-69 years and comparison men without the premutation in different decades of life, by colleagues in the UK led by Dr. Kim Cornish **[7,8]**. The primary conclusion of this study, effectively highlighted by graphs showing decreasing scores with advanced decades of life, is that age is correlated with increasingly worse performance on measures of inhibitory control, working memory and attention in PC (compared to controls). Then, a series of studies utilizing fMRI **[9,10]**, gait metrics affected by cognitive load **[11,12]**, disinhibited eye movements controlled by the cerebellum **[13]**, and a suite of structural MRI studies, including those that correlated brain with cognitive measures **[14–20]** have provided converging evidence of a dysexecutive pattern in PC with and without FXTAS.

While these cross-sectional studies revealed brain and cognition differences prior to the onset of FXTAS, the ability to determine whether cognitive changes represent prodromal features of later movement disorder relies on following individuals prospectively and longitudinally. To address this, we conducted a neuropsychological study, utilizing the Cambridge Neuropsychological Automated Battery (CANTAB), which included 64 male *FMR1* PC without FXTAS at study entry and 30 healthy controls (HC), aged 40 to 80 years. Fifty of the PC and 22 of the HC were re-assessed after an average of 2.33 years, and 37 PC and 20 HC were re-assessed a third time after an average of another 2.15 years. Results showed that changes in visual working memory, inhibitory control, and planning progress at a higher rate in PC than matched HC, and that worsening inhibitory control and planning tracked the onset of FXTAS **[21]**. Although these neuropsychological tests can reveal performance-based changes in the clinic setting, it is critically important to also understand whether executive function changes affect PC in their daily lives, and if present, whether they precede or track with the onset of FXTAS. To address these questions, the present study examined self-reported and spouse/partner-reported EF symptoms on the Behavior Rating Inventory of Executive Function-Adult Version (BRIEF-A; **[22]**)a standardized measure that captures an adult’s executive functions or self-regulation in his everyday environment, in this longitudinal cohort of PC and HC.

## 2. Materials and Methods

Our ongoing prospective longitudinal study examines trajectories of neuropsychological, motor, brain, and molecular changes in male fragile X PCs and HC with a focus on the FXTAS prodromal period and early neurodegenerative process. Participants in the present analysis include 66 PC ranging from 40-78 years (Mean=59.5) and 31 HC ages 40-75 (Mean 57.7) at baseline (visit 1). PC were recruited from throughout North America, from a variety of sources including patient referrals for clinical evaluation, the announcements by the National Fragile X Foundation, and flyers posted on social media websites dedicated to fragile X families. HC were recruited from flyers posted in the local community and on the university campus. PC visits to the research center usually entailed compensated air travel and local lodging. An important goal of the recruitment process was the inclusion of PC who do not yet have FXTAS at enrollment. During phone screening, detailed interviewing with potential participants suggested than none of the enrolled PC had FXTAS based upon criterion of evidence of either tremor or ataxia with any degree of impairment in functioning. Despite the screening, 11 of the 66 PC demonstrated at least equivocal/intermittent tremor and/or ataxia and functional interference (FXTAS Stage 2 or higher) at visit 1 after neurological examination. Overall, eighty-four participants returned for 2-5 follow up visits over a total duration of 1 to 9 years (Mean=4.6 years). Among PC, 59/69 had two or more visits, 40 had three or more, 17 had four or more, 2 had five visits. Among HC, 27/31 had two or more visits, 19 had three or more, 16 had four or more, and 2 had five visits. The primary reasons for lack of follow up visits were travel inconvenience or problems with ambulation preventing travel, busy work schedules, COVID pandemic university restrictions on in-person research, and inability to contact participants due to moves. Of the PC participants without evidence of FXTAS at baseline who had follow-up visits to date, 28 “photoconverted” and developed FXTAS (tremor and/or ataxia and functional interference). One of the PC died as a result of FXTAS complications.

The Behavior Rating Inventory of Executive Function-Adult Version (BRIEF-A; **[22]**), a standardized measure that captures views of an adult’s executive functions or self-regulation in his everyday environment, was completed by the participants and their spouses/partners completed the BRIEF-A other-report about the participant at each visit. FXTAS stage also was evaluated at each visit to track disease emergence and progression. This analysis focuses on the two broader indices of the BRIEF-A (Behavioral Regulation, BRI; and Metacognition, MCI) and the overall Global Executive Composite (GEC).

## 3. Results

There were no significant group differences at baseline (visit 1) in age, full scale IQ, education level, BRI, MCI, or GEC, with mean BRIEF-A T-scores very close to average for age in both groups (Table 1). Longitudinal mixed-effects models, however, showed a greater decline with age (both linear and quadratic) in PC compared to HC on the self-report MCI, t(134), 2.34, p=0.02, but not on the BRI or GEC (Table 2).

**Table 1.**
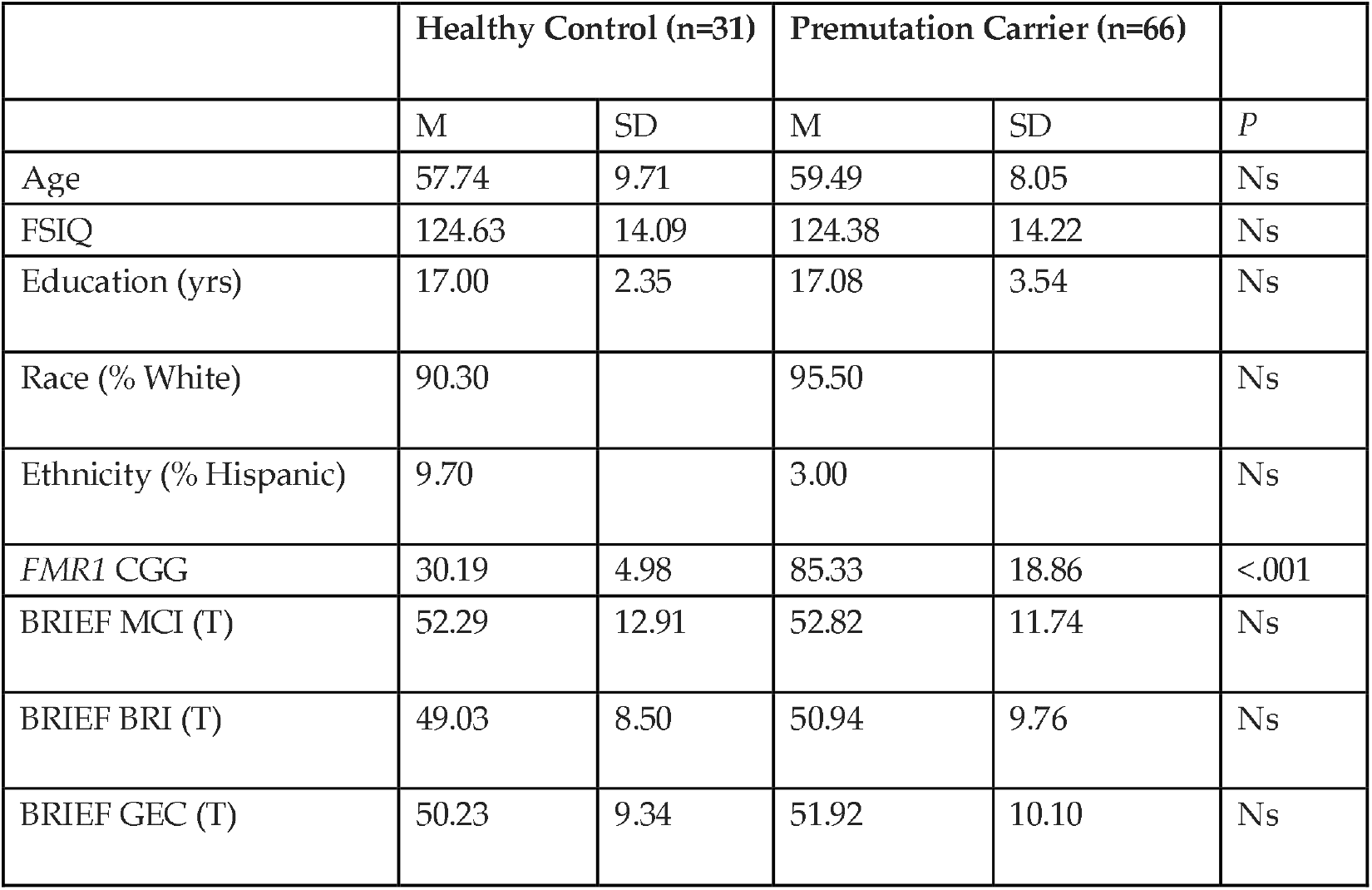
Baseline (visit 1) demographic and BRIEF-A descriptive statistics by group.

**Table 2.** Results of longitudinal mixed model regression examining the effect of age, group (PC vs. HC), and interaction of age and group on BRIEF self-report raw scores.

|  | Estimate | t | P |
| --- | --- | --- | --- |
| **Self-BRI** |  |  |  |
| Intercept | 51.09 | 11.11 | <.001 |
| Age | -1.21 | -2.67 | 0.009 |
| Age<sup>2</sup> | 0.03 | 2.75 | 0.007 |
| Group | 0.21 | 0.03 | ns |
| Age*Group | 0.12 | 0.20 | ns |
| Age<sup>2</sup>*Group | -0.00025 | -0.02 | ns |

| Self-MI | Estimate | t | P |
| --- | --- | --- | --- |
| Intercept | 73.03 | 11.08 | <.001 |
| Age | -2.05 | -3.34 | 0.001 |
| Age<sup>2</sup> | 0.05 | 3.72 | <.001 |
| Group | -15.12 | -1.66 | ns |
| Age*Group | 1.94 | 2.30 | 0.023 |
| Age<sup>2</sup>*Group | -0.04 | -2.34 | 0.021 |

In premutation carriers, conversion to FXTAS based upon advancing FXTAS Stage in carriers of older age was associated with worsening BRI, t(83)=2.59, p=.01, MI, t(81)=2.03, p=.04 and GEC, t(83)=2.17, p=.03 (Table 3).

**Table 3.**
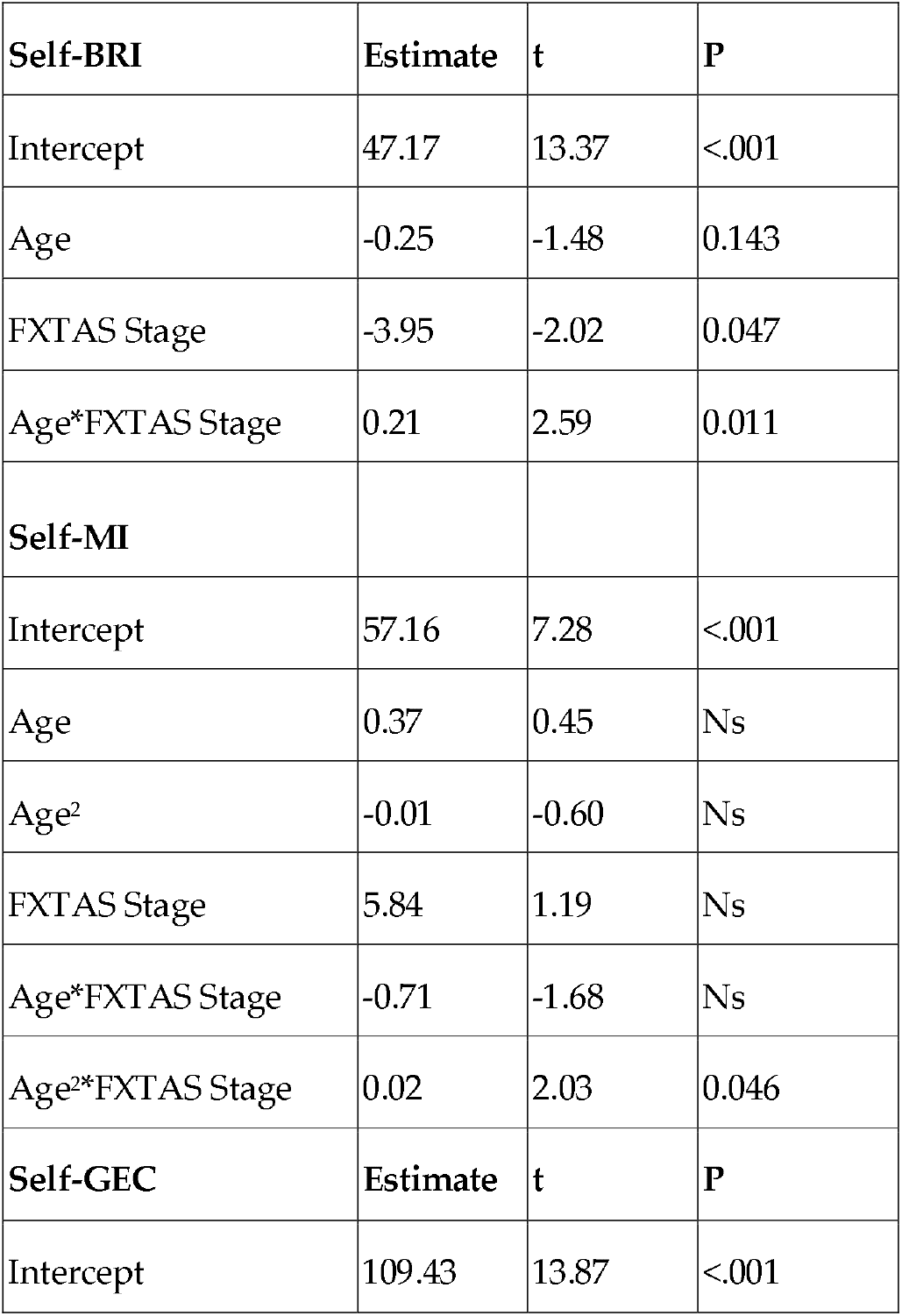
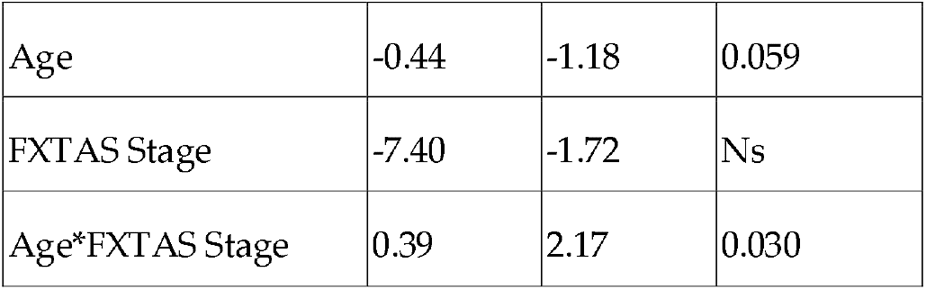
Results of longitudinal mixed model regression examining the effect of age, FXTAS Stage, and interaction of age and FXTAS stage on BRIEF self-report raw scores (PC only).

Survival analyses predicting the probability of converting to FXTAS (stage 2 or higher), utilizing age and BRIEF self report-revealed that baseline measures of age (OR = 1.09, SE = .046, p = .048), higher BRIEF GEC (OR = 1.03, SE = .014, p = .047), and MI (OR = 1.06, SE = .024, p = .020), but not BRI, were significantly associated with probability of later FXTAS conversion.

For spouse/partner report BRIEF-A, no difference in trajectory was found between groups, however FXTAS conversion was associated with worsening GEC, t(77) = 2.07, p=.04, and this effect approached significance for the MI (Table 4). Agreement between PCs and spouse/partners was positive and statistically significant, but the magnitude of agreement was modest (e.g., PC GEC Intraclass Correlation Coefficient=.55.; HC GEC Intraclass Correlation Coefficient =.79).

**Table 4.** Results of longitudinal mixed model regression examining the effect of age, FXTAS Stage, and interaction of age and FXTAS stage on BRIEF spouse/partner-report raw scores (PC only).

| Spouse/Partner-MI | Estimate | t | P |
| --- | --- | --- | --- |
| Intercept | 68.15 | 11.31 | <.001 |
| Age | -0.57 | -1.91 | Ns |
| FXTAS Stage | -4.73 | -1.35 | Ns |
| Age*FXTAS Stage | 0.30 | 1.95 | 0.05 |
| **Spouse/Partner-GEC** |  |  |  |
| Intercept | 115.64 | 11.22 | <.001 |
| Age | -0.70 | -1.41 | ns |
| FXTAS Stage | -8.58 | -1.59 | ns |
| Age*FXTAS Stage | 0.49 | 2.07 | 0.04 |

## 4. Discussion

This study examined the ratings of executive function problems of adult male *FMR1* premutation carriers (PC) and a well-matched group of healthy men without the premutation (HC) in a prospective longitudinal study. A unique aspect of the project is the focus on enrollment of PC before the onset of FXTAS in order to study its prodrome and earliest indicators of disease onset and progression. At baseline, with a mean age of about 49 years, PC and HC reported normal rates of executive function challenges with nearly identical standard scores on the BRIEF-A. Over time, however, PC reported a greater decline in metacognition (composed of self-initiation, working memory, organization, task monitoring) with age than HC, but not a significantly worse decline in behavioral regulation. Declines in both metacognition and behavioral regulation were associated with onset and progression of FXTAS in those that developed the disease over time. Interestingly, spouses/partners were able to notice and report changes in EF among those who progressed to FXTAS but seemed less able to detect the EF changes in PC generally. The survival analysis provided preliminary evidence that the PC with relative weaknesses in metacognition are at higher risk for later neurodegeneration. The results also seem to indicate that PC are aware of changes in their metacognition before it can be recognized in outward behavior by others.

These results lend support to prior cross-sectional studies suggesting that EF changes precede FXTAS and may be considered a prodromal feature of the movement disorder, similar to what has been observed in Parkinson’s disease (PD). In two large prospective cohort studies of community dwelling elders, men who later developed idiopathic PD showed a faster decline in executive function in the prediagnostic period, followed by a steeper decline after diagnosis **[23]**. Indeed, parkinsonism is a common feature of FXTAS and many individuals with FXTAS are misdiagnosed with PD **[24–26]**. If PC experience EF changes before FXTAS develops and are an aspect of the earliest manifestations of disease, it is possible that measures could be developed that are especially sensitive to the specific EF problems that precede the onset of the movement disorder. This is a challenge, however, given that normal aging is associated with changes in EF, and such tools would need to separate FXTAS-associated executive changes from normal aging decline. Studies are needed to compare and contrast the sensitivity and specificity of predictive clinical measures and biomarkers in PC at risk for FXTAS. This is likely to require larger samples through combined efforts of investigators in consortia and standardized measures and procedures. With some targeted treatment trials on the horizon **[27–29]**, this work will be critical in order to identify the most optimal measures to identify PC most at risk and to track response to treatment.

What are the brain mechanisms underlying motor and executive function decline in male carriers, and how are they connected to FXTAS? A growing body of research suggests that deterioration of frontal-cerebellar circuits is especially sensitive to the *FMR1* premutation and appear to impact both control of movement (ataxia and tremor) and executive control of cognition (e.g., working memory) in PC **[9,19,30–32]**. For example, we previously showed that compared with HC, PC with and without FXTAS performing a verbal working memory EF task showed reduced activation in the right ventral inferior frontal cortex and left premotor/dorsal inferior frontal cortex **[9]**. In a cross-sectional structural brain imaging study of 322 males from age 8 to 81, we discovered that compared to age-matched HC, PC without FXTAS showed significantly accelerated age-related volume decreases in the cerebellum and brainstem and increased ventricular volume **[18]**.

One of the limitations of the study is the unequal sample sizes, with fewer HC than PC. The project was designed in this way to maximize characterization of PC trajectories of change and examine predictors of phenoconversion in PC, given limited resources including domestic travel and imaging costs. Another limitation is the exclusion of female PC, and as such, the study can only be generalized to males. The reason for the focus on males is that the project was designed expressly to study the prodrome of FXTAS and factors that predict onset of disease. Given that the PC is an X-linked condition, and that only about 8-16% of women develop FXTAS **[33]**, we chose to focus on men who are much more likely to phenoconvert during the study period. Whether EF problems are associated with neurodegenerative or other health changes in female PC is an open question. Also, the results of this study may not generalize to the full population of male PC because this project included a predominantly white, relatively affluent group of men who were able to travel and take time away from work. In the future, we plan to examine self-reported and neuropsychological test-related changes in EF in concert with structural MRI changes over time in PC vs HC to obtain a more integrated picture of how neurodegeneration unfolds and determine which metrics are most useful to collect in intervention studies including clinical trials. Nevertheless, it is clear from this and prior work that EF (and especially EF change) is a critical cognitive construct to assess in fragile X PC.

## Data Availability

All data produced in the present study are available upon reasonable request to the authors.

## Author Contributions

Conceptualization, D.H. and S.R.; methodology, D.H., E.F., F.T., and S.R.; formal analysis, E.F.; investigation, D.H., S.R., A.S., R.H., and G.E.; data curation, G.E. and K.M.R.; writing—original draft preparation, D.H.; writing— review and editing, X.X.; supervision, D.H. and S.R.; project administration, D.H and G.E.; funding acquisition, D.H. and S.R. All authors have read and agreed to the published version of the manuscript.

## Institutional Review Board Statement

The study was conducted in accordance with the Declaration of Helsinki and approved by the Institutional Review Board of the University of California Davis (protocol code 473010, approval 7/11/2013).

## Informed Consent Statement

Informed consent was obtained from all subjects involved in the study.

## Data Availability Statement

The datasets used and/or analyzed during the current study are available from the corresponding author on reasonable request.

## Acknowledgments

We thank the participants and their families for their effort and dedication to this research. We also thank Drs. Corrisa Jacomini and Sundas Pasha for neuropsychological examinations of participants.

## References

1. Dombrowski, C.; Lévesque, S.; Morel, M.L.; Rouillard, P.; Morgan, K.; Rousseau, F. Premutation and Intermediate-Size FMR1 Alleles in 10572 Males from the General Population: Loss of an AGG Interruption Is a Late Event in the Generation of Fragile X Syndrome Alleles. Hum Mol Genet 2002, 11, 371–378, doi:10.1093/hmg/11.4.371.

2. Tassone, F.; Iong, K.P.; Tong, T.-H.; Lo, J.; Gane, L.W.; Berry-Kravis, E.; Nguyen, D.; Mu, L.Y.; Laffin, J.; Bailey, D.B.; et al. FMR1 CGG Allele Size and Prevalence Ascertained through Newborn Screening in the United States. Genome Med 2012, 4, 100, doi:10.1186/gm401.

3. Bacalman, S.; Farzin, F.; Bourgeois, J.A.; Cogswell, J.; Goodlin-Jones, B.L.; Gane, L.W.; Grigsby, J.; Leehey, M.A.; Tassone, F.; Hagerman, R.J. Psychiatric Phenotype of the Fragile X-Associated Tremor/Ataxia Syndrome (FXTAS) in Males: Newly Described Fronto-Subcortical Dementia. J Clin Psychiatry 2006, 67, 87–94, doi:10.4088/jcp.v67n0112.

4. Grigsby, J.; Brega, A.G.; Jacquemont, S.; Loesch, D.Z.; Leehey, M.A.; Goodrich, G.K.; Hagerman, R.J.; Epstein, J.; Wilson, R.; Cogswell, J.B.; et al. Impairment in the Cognitive Functioning of Men with Fragile X-Associated Tremor/Ataxia Syndrome (FXTAS). J Neurol Sci 2006, 248, 227–233, doi:10.1016/j.jns.2006.05.016.

5. Grigsby, J.; Brega, A.G.; Engle, K.; Leehey, M.A.; Hagerman, R.J.; Tassone, F.; Hessl, D.; Hagerman, P.J.; Cogswell, J.B.; Bennett, R.E.; et al. Cognitive Profile of Fragile X Premutation Carriers with and without Fragile X-Associated Tremor/Ataxia Syndrome. Neuropsychology 2008, 22, 48–60, doi:10.1037/0894-4105.22.1.48.

6. Grigsby, J.; Brega, A.G.; Leehey, M.A.; Goodrich, G.K.; Jacquemont, S.; Loesch, D.Z.; Cogswell, J.B.; Epstein, J.; Wilson, R.; Jardini, T.; et al. Impairment of Executive Cognitive Functioning in Males with Fragile X-Associated Tremor/Ataxia Syndrome. Mov Disord 2007, 22, 645–650, doi:10.1002/mds.21359.

7. Cornish, K.M.; Kogan, C.S.; Li, L.; Turk, J.; Jacquemont, S.; Hagerman, R.J. Lifespan Changes in Working Memory in Fragile X Premutation Males. Brain Cogn 2009, 69, 551–558, doi:10.1016/j.bandc.2008.11.006.

8. Cornish, K.M.; Li, L.; Kogan, C.S.; Jacquemont, S.; Turk, J.; Dalton, A.; Hagerman, R.J.; Hagerman, P.J. Age-Dependent Cognitive Changes in Carriers of the Fragile X Syndrome. Cortex 2008, 44, 628–636, doi:10.1016/j.cortex.2006.11.002.

9. Hashimoto, R.; Backer, K.C.; Tassone, F.; Hagerman, R.J.; Rivera, S.M. An FMRI Study of the Prefrontal Activity during the Performance of a Working Memory Task in Premutation Carriers of the Fragile X Mental Retardation 1 Gene with and without Fragile X-Associated Tremor/Ataxia Syndrome (FXTAS). J Psychiatr Res 2011, 45, 36–43, doi:10.1016/j.jpsychires.2010.04.030.

10. Wang, J.M.; Koldewyn, K.; Hashimoto, R.-I.; Schneider, A.; Le, L.; Tassone, F.; Cheung, K.; Hagerman, P.; Hessl, D.; Rivera, S.M. Male Carriers of the FMR1 Premutation Show Altered Hippocampal-Prefrontal Function during Memory Encoding. Front Hum Neurosci 2012, 6, 297, doi:10.3389/fnhum.2012.00297.

11. O’Keefe, J.A.; Guan, J.; Robertson, E.; Biskis, A.; Joyce, J.; Ouyang, B.; Liu, Y.; Carnes, D.; Purcell, N.; Berry-Kravis, E.; et al. The Effects of Dual Task Cognitive Interference and Fast-Paced Walking on Gait, Turns, and Falls in Men and Women with FXTAS. Cerebellum 2021, 20, 212–221, doi:10.1007/s12311-020-01199-3.

12. O’Keefe, J.A.; Robertson, E.E.; Ouyang, B.; Carns, D.; McAsey, A.; Liu, Y.; Swanson, M.; Bernard, B.; Berry-Kravis, E.; Hall, D.A. Cognitive Function Impacts Gait, Functional Mobility and Falls in Fragile X-Associated Tremor/Ataxia Syndrome. Gait Posture 2018, 66, 288–293, doi:10.1016/j.gaitpost.2018.09.005.

13. Wong, L.M.; Goodrich-Hunsaker, N.J.; McLennan, Y.; Tassone, F.; Zhang, M.; Rivera, S.M.; Simon, T.J. Eye Movements Reveal Impaired Inhibitory Control in Adult Male Fragile X Premutation Carriers Asymptomatic for FXTAS. Neuropsychology 2014, 28, 571–584, doi:10.1037/neu0000066.

14. Cvejic, R.C.; Hocking, D.R.; Wen, W.; Georgiou-Karistianis, N.; Cornish, K.M.; Godler, D.E.; Rogers, C.; Trollor, J.N. Reduced Caudate Volume and Cognitive Slowing in Men at Risk of Fragile X-Associated Tremor Ataxia Syndrome. Brain Imaging Behav 2019, 13, 1128–1134, doi:10.1007/s11682-018-9928-7.

15. Filley, C.M. Fragile X Tremor Ataxia Syndrome and White Matter Dementia. Clin Neuropsychol 2016, 30, 901–912, doi:10.1080/13854046.2016.1165805.

16. Hippolyte, L.; Battistella, G.; Perrin, A.G.; Fornari, E.; Cornish, K.M.; Beckmann, J.S.; Niederhauser, J.; Vingerhoets, F.J.G.; Draganski, B.; Maeder, P.; et al. Investigation of Memory, Executive Functions, and Anatomic Correlates in Asymptomatic FMR1 Premutation Carriers. Neurobiol Aging 2014, 35, 1939–1946, doi:10.1016/j.neurobiolaging.2014.01.150.

17. Shelton, A.L.; Cornish, K.M.; Godler, D.; Bui, Q.M.; Kolbe, S.; Fielding, J. White Matter Microstructure, Cognition, and Molecular Markers in Fragile X Premutation Females. Neurology 2017, 88, 2080–2088, doi:10.1212/WNL.0000000000003979.

18. Wang, J.Y.; Hessl, D.; Hagerman, R.J.; Simon, T.J.; Tassone, F.; Ferrer, E.; Rivera, S.M. Abnormal Trajectories in Cerebellum and Brainstem Volumes in Carriers of the Fragile X Premutation. Neurobiol Aging 2017, 55, 11–19, doi:10.1016/j.neurobiolaging.2017.03.018.

19. Wang, J.Y.; Grigsby, J.; Placido, D.; Wei, H.; Tassone, F.; Kim, K.; Hessl, D.; Rivera, S.M.; Hagerman, R.J. Clinical and Molecular Correlates of Abnormal Changes in the Cerebellum and Globus Pallidus in Fragile X Premutation. Front Neurol 2022, 13, 797649, doi:10.3389/fneur.2022.797649.

20. Yang, J.-C.; Chan, S.-H.; Khan, S.; Schneider, A.; Nanakul, R.; Teichholtz, S.; Niu, Y.-Q.; Seritan, A.; Tassone, F.; Grigsby, J.; et al. Neural Substrates of Executive Dysfunction in Fragile X-Associated Tremor/Ataxia Syndrome (FXTAS): A Brain Potential Study. Cereb Cortex 2013, 23, 2657–2666, doi:10.1093/cercor/bhs251.

21. Famula, J.; Ferrer, E.; Hagerman, R.J.; Tassone, F.; Schneider, A.; Rivera, S.M.; Hessl, D. Neuropsychological Changes in FMR1 Premutation Carriers and Onset of Fragile X-Associated Tremor/Ataxia Syndrome. J Neurodev Disord 2022, 14, 23, doi:10.1186/s11689-022-09436-y.

22. Roth, R.; Isquith, P.; Gioia, G. BRIEF-A Behavior Rating Inventory of Executive Function-Adult Version, Publication Manual; Psychological Assessment Resources, Inc, 2005;

23. Bock, M.A.; Vittinghoff, E.; Bahorik, A.L.; Leng, Y.; Fink, H.; Yaffe, K. Cognitive and Functional Trajectories in Older Adults With Prediagnostic Parkinson Disease. Neurology 2023, 100, e1386–e1394, doi:10.1212/WNL.0000000000206762.

24. Niu, Y.-Q.; Yang, J.-C.; Hall, D.A.; Leehey, M.A.; Tassone, F.; Olichney, J.M.; Hagerman, R.J.; Zhang, L. Parkinsonism in Fragile X-Associated Tremor/Ataxia Syndrome (FXTAS): Revisited. Parkinsonism Relat Disord 2014, 20, 456–459, doi:10.1016/j.parkreldis.2014.01.006.

25. Kartanou, C.; Seferiadi, M.; Pomoni, S.; Potagas, C.; Sofocleous, C.; Traeger-Synodinos, J.; Stefanis, L.; Panas, M.; Koutsis, G.; Karadima, G. Screening for the FMR1 Premutation in Greek Patients with Late-Onset Movement Disorders. Parkinsonism Relat Disord 2023, 107, 105253, doi:10.1016/j.parkreldis.2022.105253.

26. Hall, D.A.; Howard, K.; Hagerman, R.; Leehey, M.A. Parkinsonism in FMR1 Premutation Carriers May Be Indistinguishable from Parkinson Disease. Parkinsonism Relat Disord 2009, 15, 156–159, doi:10.1016/j.parkreldis.2008.04.037.

27. Kong, H.E.; Zhao, J.; Xu, S.; Jin, P.; Jin, Y. Fragile X-Associated Tremor/Ataxia Syndrome: From Molecular Pathogenesis to Development of Therapeutics. Frontiers in Cellular Neuroscience 2017, 11.

28. Wang, J.Y.; Trivedi, A.M.; Carrillo, N.R.; Yang, J.; Schneider, A.; Giulivi, C.; Adams, P.; Tassone, F.; Kim, K.; Rivera, S.M.; et al. Open-Label Allopregnanolone Treatment of Men with Fragile X-Associated Tremor/Ataxia Syndrome. Neurotherapeutics 2017, 14, 1073–1083, doi:10.1007/s13311-017-0555-6.

29. Napoli, E.; Schneider, A.; Wang, J.Y.; Trivedi, A.; Carrillo, N.R.; Tassone, F.; Rogawski, M.; Hagerman, R.J.; Giulivi, C. Allopregnanolone Treatment Improves Plasma Metabolomic Profile Associated with GABA Metabolism in Fragile X-Associated Tremor/Ataxia Syndrome: A Pilot Study. Mol Neurobiol 2019, 56, 3702–3713, doi:10.1007/s12035-018-1330-3.

30. Shelton, A.L.; Wang, J.Y.; Fourie, E.; Tassone, F.; Chen, A.; Frizzi, L.; Hagerman, R.J.; Ferrer, E.; Hessl, D.; Rivera, S.M. Middle Cerebellar Peduncle Width-A Novel MRI Biomarker for FXTAS? Front Neurosci 2018, 12, 379, doi:10.3389/fnins.2018.00379.

31. Brown, S.S.G.; Basu, S.; Whalley, H.C.; Kind, P.C.; Stanfield, A.C. Age-Related Functional Brain Changes in FMR1 Premutation Carriers. Neuroimage Clin 2018, 17, 761–767, doi:10.1016/j.nicl.2017.12.016.

32. Wang, J.Y.; Hessl, D.; Schneider, A.; Tassone, F.; Hagerman, R.J.; Rivera, S.M. Fragile X-Associated Tremor/Ataxia Syndrome: Influence of the FMR1 Gene on Motor Fiber Tracts in Males with Normal and Premutation Alleles. JAMA Neurol 2013, 70, 1022–1029, doi:10.1001/jamaneurol.2013.2934.

33. Hagerman, R.; Hagerman, P. Advances in Clinical and Molecular Understanding of the FMR1 Premutation and Fragile X-Associated Tremor/Ataxia Syndrome. The Lancet Neurology 2013, 12, 786–798, doi:10.1016/S1474-4422(13)70125-X.

